# Global and regional prevalence and outcomes of COVID-19 in people living with HIV: A cutting-edge systematic review and meta-analysis

**DOI:** 10.1101/2021.07.12.21260361

**Authors:** Reynie Purnama Raya, Ami Kamila, Jaber S Alqahtani, Ahmed M Hjazi, Amy Li, Tope Oyelade

**Author notes:** Corresponding Author – Tope Oyelade.

## Abstract

**Background:** The relationship between HIV infection and COVID-19 clinical outcome is uncertain, with conflicting data and hypotheses. We aimed to assess the prevalence and risk of severe COVID-19 and death in people living with HIV (PLWH) on the global and continental level.

**Methods:** Electronic databases were systematically searched in July 2021. Studies were screened and then extracted following the Preferred Reporting Items for Systematic Reviews and Meta-Analyses guidelines. Narratives were synthesised and data pooled for global and continental prevalence and relative risk of severity and mortality in HIV-infected COVID-19 patients using random-effect model. Risk of bias was assessed using the Newcastle-Ottawa score, Egger’s test and presented as funnel plots.

**Results:** A total of 46 studies were included involving 18,034,947 COVID-19 cases of which 31,269 were PLWH. The global prevalence of PLWH with SARS-CoV-2 infection was 1% (95% CI = 0.9% -1.1%) with the highest prevalence observed in sub-Saharan Africa. The relative risk (RR) of COVID-19 severity was significant only in Africa (RR, 95% CI = 1.14, 1.08 – 1.24) while risk of COVID-19 mortality was 1.53% (95% CI = 1.45 – 2.03) globally. The prevalence of PLWH in COVID-19 cases was significantly low, and the calculated global risk ratio show that HIV infection may be linked with increased COVID-19 death. The between-studies heterogeneity was significantly high while risk of publication bias was not significant.

**Conclusion:** There is low prevalence of HIV-SARS-CoV-2 co-infection. HIV infection was linked with severe COVID-19 in Africa and increased risk of death globally.

## Introduction

The 2019 coronavirus (COVID-19) pandemic caused by SARS-CoV-2 remains a global public health challenge that has affected over 186 million people and caused over 4 million deaths globally (1). While most cases of COVID-19 are clinically mild or asymptomatic, older age and certain underlying illness such as cardiovascular, respiratory, and digestive diseases have been reported to increase risk of severe COVID-19 cases or death (2-4). Such comorbidities are associated with an increased fatality rate and present a challenge for intensive care management of COVID-19 patients (5, 6).

Human Immunodeficiency Virus (HIV) belongs to a genus of zoonotic lentiviruses that cause acute immune-deficiency syndrome (AIDS) (7). Data from the Joint United Nations Programme on HIV/AIDS (UNAIDS) puts the number of people living with HIV (PLWH) at 38 million globally with 1.5 million new infections in 2020 and about 6 million not knowing their HIV infection status (8). Accordingly, the number of PLWH is projected to increase due to treatment availability and the associated reduction in AIDS-related deaths (9).

HIV is associated with a dysregulation of the immune system which predisposes patients to opportunistic infectious diseases (10). Indeed, most HIV-related death have been linked to secondary infections and abnormal inflammatory response resulting from AIDS (11) This is especially so in patients with uncontrolled HIV replication, high viral load and low CD4/CD8 count. Giving the immune-compromised state of most PLWH and increased possibility of secondary dysfunctions, an increased risk of infection, severity, and death due to COVID-19 may be expected. However, attenuated immune response may also protect against cytokine release storm and corresponding acute respiratory distress syndrome (ARDS) linked with SARS-CoV-2 infection (12). Hence, the association between HIV infection and COVID-19 remain unclear with sparse and conflicting reports (13, 14). Aside from the heterogeneity from difference in epidemiology of HIV between countries and continents, variability exist in treatment, management, and behaviour of PLWH. These amongst other factors, determines the rate of spread as well as the availability and uptake of HIV preventative and treatment measures (15). This is further exacerbated by the disruption to clinical care of various chronic diseases due to the diversion of medical resources to manage the burgeoning COVID-19 cases around the world (16). This review aimed to provide an updated insight into the global and continental prevalence, potential risk of severe illness and death due to COVID-19 in PLWH by conducting a meta-analysis grouped by continents.

## Methods

The protocol of this systematic review was registered prospectively to PROSPERO (CRD42021264151). Following the Preferred Reporting in Systematic Reviews and Meta-Analyses (PRISMA) guidelines (17), Medline and Embase databases were searched on 2^nd^ of July 2021 using keywords and MeSH terms (Fig. S1). Further, a search of pre-printed papers was performed because of the rapidly developing nature of the topic. Studies retrieved from the search were imported into EndNote software and the duplicates removed. The resulting duplicated-free studies were then uploaded to Rayyan software and title, abstract and whole text screening was carried out (RPR, AK). Manual reference search was also performed to cover papers that were not covered by the search strategy (Fig. 1).

**Figure 1.**
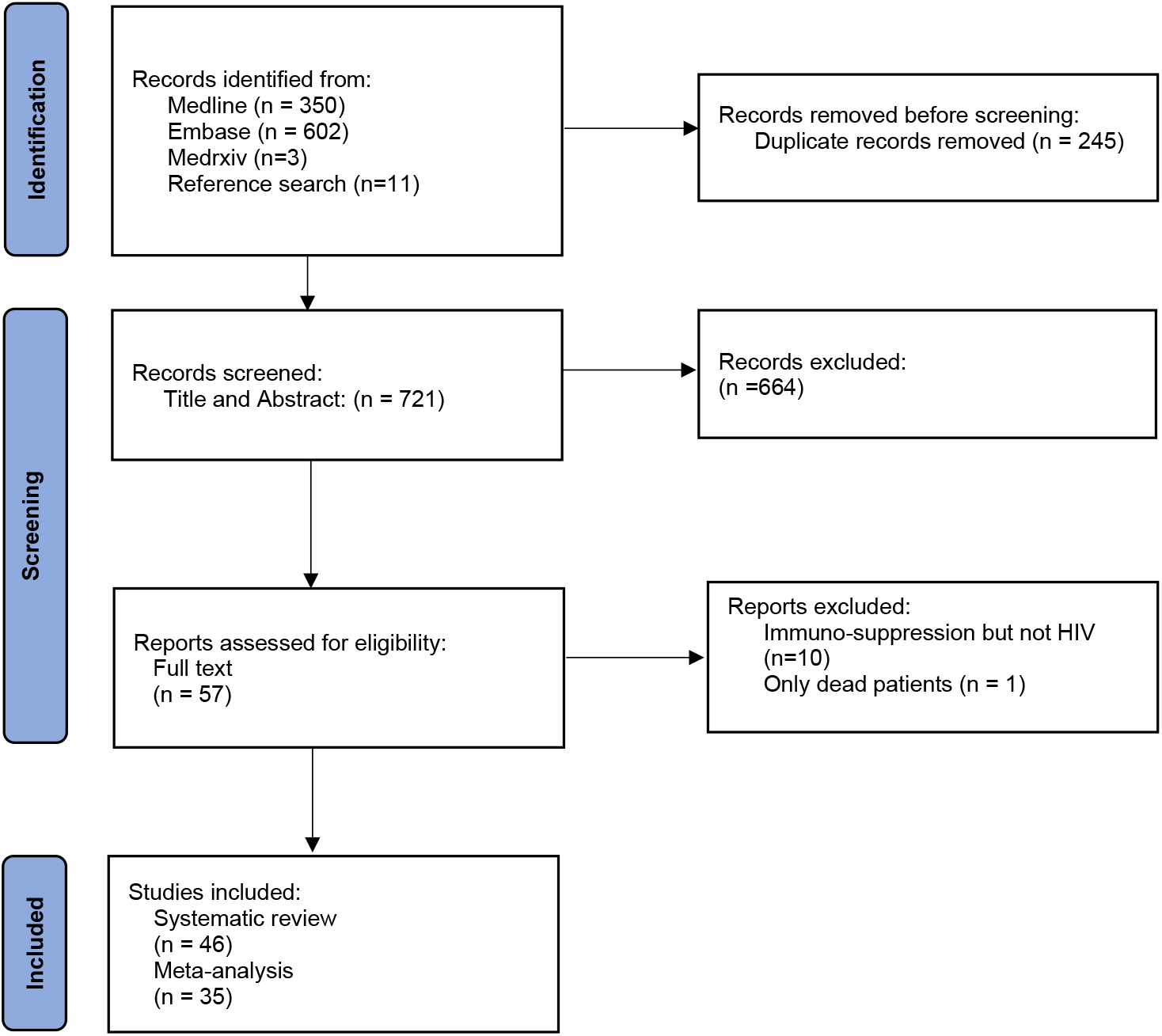
Global and regional prevalence and outcomes of COVID-19 in people living with HIV: A systematic review and meta-analysis according to the Preferred Reporting for Systematic Reviews and Meta-analyses diagram.

### Inclusion and exclusion criteria

The inclusion/exclusion of studies included followed the PECO (Population, Exposure, Comparison and Outcome) model and only involve studies that presents the clinical characteristics and/or composite endpoints of COVID-19 in people living with HIV (PLWH). Studies that combined HIV and other immunosuppressive diseases and conditions (cancer, congenital, or medically induced), non-English language, reviews, case reports, qualitative studies, editorials and were excluded.

### Data collections

Two authors (RPR, AK) independently screened titles and abstracts of potentially eligible studies and any disagreements were resolved by discussing with a third reviewer (TO). Full texts of the studies that were included based on the abstract were fully read and assess against the inclusion/exclusion criteria.

### Data extraction and analysis

The authors and year of publication, study design and period, country of study, sample size, number of PLWH and non-HIV COVID-19 patients as well as their clinical outcomes and biometrics data were extracted into a table. Clinical outcomes identified were severity of the disease and death. Severe COVID-19 was described as prolonged hospital stay, ICU admission and requirement of mechanical ventilation (MV). Analyses were performed using Stata/MP 17; prevalence was calculated by ”metaprop” procedure. Forest plots were used to present the pooled prevalence of PLWH in COVID-19 cases grouped by continents of studies. Continent-grouped effect sizes (95% Confidence Intervals, CIs) and the test results of between-studies heterogeneity (I^2^ Statistic, p value) were also computed. The ‘metan’ procedure was used to assess risk of severity and mortality in PLWH-COVID-19 patients compared with general population in the included studies and the risk ratios were grouped by continents to further assess the intercontinental variation in these risks.

### Quality assessment

A modified version of the Newcastle-Ottawa Score (NOS) was used to assess the risk of bias in the included studies (18). This includes 3 domains and 9 questions scored accordingly with a star if satisfied. The selection domain assessed the randomness, multicentre involvement in selection of study population as well as sample size. Sample size of ≥ 100 was decided based on previous studies’ estimate of ∼1% prevalence of HIV infection in COVID-19 cases (19, 20). The standard ascertainment of COVID-19 and HIV were also assessed against the WHO guideline (21). Finally, the follow up time (≥2 weeks), mode of outcome confirmation and whether all patients were accounted for were assessed (Table S1). Studies with ≥5 stars (> 50%) were considered unbiased. To further assess publication bias in the studies pooled for prevalence and the risk of severity and mortality in PLWH-COVID-19, funnel plots and Egger test were computed using ‘metafunnel’ and ‘metabias’ procedures respectively in STATA. Statistical significance was set at 95% (p < 0.05).

## Results

The systematic search of databases including preprints and reference search generated an initial total of 966 studies including 245 duplicates to give a total of 721 studies. Initial title and abstract screening led in the exclusion of 652 studies followed by full-text review and further exclusion of 11 studies to give a total of 46 studies which satisfy the inclusion/exclusion criteria (Fig. 1).

### General description of studies included

The 46 studies in the systematic review included 18,190,619 COVID-19 patients of which 36,633 (0.2%) are PLWH. The sample sizes of the included studies ranged from 20 to 17,282,905 with data from 15 countries across 4 continents. Overall, 30 of the studies were retrospective with 10 prospective, 4 case series, 2 descriptive (Table 1). The risk of bias assessment showed low bias in the included studies with 87% (40/46) of the studies below the bias threshold (Table S1).

**Table 1 -.** General characteristics of included studies.

| Study name (year) | Country | Type of study | Sample size (M; F; T) | Age Mean±SD or<br>median, range | PLWH | PLWH<br>survive | PLWH<br>non-<br>survive | PLWH<br>Severe | PLWH Non-<br>severe |
| --- | --- | --- | --- | --- | --- | --- | --- | --- | --- |
| Bhaskaran et al (2021) (22) | UK | Retrospective | 17282905 (NR) | NR | 27480 | 27455 | 25 | NR | NR |
| Di Biagio et al (2020) (23) | Italy | Prospective | 69(NR; All HIV) | NR | 69 | 62 | 7 | 4 | 58 |
| Borobia et al (2020) (24) | Spain | Retrospective | 2226 (M=1074; F=1152) | 61 (IQR 46-78) | 13 | 9 | 4 | NR | NR |
| Boulle et al (2020) (25) | South Africa | Retrospective | 22308(NR) | (NR) | 3978 | 3863 | 115 | 601 | 3262 |
| Byrd et al (2020) (26) | USA | Case series | 27 (M=20; F=6; T=1) | NR | 27 | 26 | 1 | NR | NR |
| Canevelli et al (2020) (27) | Italy | Retrospective | 2687 (NR) | NR | 6 | NR | 6 | NR | NR |
| Ceballos et al (2021) (28) | Chile | Prospective | 18321(M=10300; F=8021) | NR | 36 | 31 | 5 | 11 | 25 |
| Collins et al (2020) (29) | USA | Case series | 530 (NR) | NR | 20 | 17 | 3 | 3 | 17 |
| Del Amo et al (2020) (30) | Spain | Prospective | 236(M=204; F=32; All HIV) | NR | 236 | 216 | 20 | 15 | 221 |
| Docherty et al (2020) (31) | UK | Prospective | 20133 (M=12068; F=8065) | 73 (IQR 58-62) | 83 | 37 | 23 | NR | NR |
| Erinoso et al (2020) (32) | Nigeria | Retrospective | 632 (M=385, F=247) | 40.1 (SD±13.9) | 3 | NR | NR | NR | NR |
| Etienne et al (2020) (33) | France | Prospective | 54(M=33; F=21; All PLWH) | 54 (range 47–60) | 54 | 53 | 1 | 19 | 35 |
| Garetti et al (2020) (34) | UK | Prospective | 47592 (NR) | NR | 122 | 75 | 30 | NR | NR |
| Gervasoni et al (2020) (35) | Italy | Retrospective | 47 (M=36; F=11) | 51 ± 11 | 47 | 45 | 2 | 2 | 34 |
| Geteneh et al (2021) (36) | Ethiopia | Retrospective | 372(M=279; F=93) | 30 (5-85) | 6 | 5 | 1 | 1 | 5 |
| Gudipati et al (2020) (37) | USA | Case series | 7372 (NR) | NR | 14 | 11 | 3 | 2 | 12 |
| Hadi et al (2020) (38) | USA | Retrospective | 50167(NR) | NR | 404 | 384 | 20 | 78 | 326 |
| Harter et al (2020) (39) | Germany | Retrospective | 33 (M=30; F=3) | 48 (range 26-82) | 33 | 29 | 3 | 8 | 25 |
| Ho et al (2021) (40) | USA | Retrospective | 93(M=67; F=23, T=3; All PLWH) | 58 (range 52–65) | 93 | 74 | 19 | 19 | 74 |
| Huang et al (2020) (41) | China | Retrospective | 50368(NR) | NR | 35 | 33 | 2 | 15 | 20 |
| Inciarte et al (2020) (42) | Spain | Prospective | 5683 (NR) | NR | 53 | 51 | 2 | 10 | 43 |
| Isernia et al (2020) (43) | French | Case series | 390 (NR) | NR | 30 | 24 | 2 | 4 | 24 |
| Izquierdo et al (2020) (44) | Spain | Retrospective | 10504 (M=5519; F=4984) | 58.2±19.7 | 34 | NR | NR | 1 | 33 |
| Karim et al (2020) (45) | South Africa | Retrospective | 124 (M=30; F=94) | 45 (IQR, 35.0-57.4) | 55 | NR | NR | 16 | 39 |
| Kirenga et al (2020) (46) | Uganda | Prospective | 56 (M=38; F=18) | 34.2 ±15.5 | 4 | 4 | 0 | NR | NR |
| Liu et al (2020) (47) | China | Retrospective | 20 (M=5; F=15) | 46.5 (IQR, 39.3 - 50.5) | 20 | 19 | 1 | 3 | 17 |
| Maggiolo et al (2021) (48) | Italy | Prospective | 55(M=44; F=11; All PLWH) | 54 (49-58) | 55 | 51 | 4 | 11 | 44 |
| Migisha et al (2020) (49) | Uganda | Retrospective | 54(M=34; F=20) | NR | 2 | 2 | 0 | 0 | 2 |
| Miyashita & Kuno (2021) (50) | USA | Retrospective | 8912(NR) | NR | 161 | 138 | 23 | 36 | 125 |
| Nachega et al (2020) (51) | Congo | Retrospective | 766(M=500; F=262; Unknown=4) | 34 ±4.5 | 12 | 10 | 2 | 3 | 9 |
| Ombajo et al (2020) (52) | Kenya | Retrospective | 787 (M=505; F=282) | 43 (range 0-109) | 53 | 42 | 11 | NR | NR |
| Parker et al (2020) (53) | South Africa | Retrospective | 113(M=44; F=69) | NR | 24 | 18 | 6 | 5 | 19 |
| Pujari et al (2021) (54) | India | Retrospective | 86 (M=66; F=20; all PLWH) | 45 ±52.3 | 86 | 80 | 6 | 17 | 69 |
| Rodriguez-Gonzalez et al (2021) (55) | Spain | Retrospective | 1255 (M=725; F=530) | 65 (range 51–77) | 12 | 9 | 3 | 1 | 11 |
| Rodriguez-Molinero et al (2020) (56) | Spain | Prospective | 418 (M=238; F=180) | 65.4 ± 16.6 | 3 | 2 | 1 | 3 | 0 |
| Shalev et al (2020) (57) | USA | Retrospective | 2159(NR) | NR | 31 | 23 | 8 | 2 | 29 |
| Shi et al (2020) (58) | China | Retrospective | 134 (M=65; F=69) | 46 (IQR, 34-58) | 1 | 1 | 0 | 0 | 1 |
| Sigel et al (2020) (59) | USA | Retrospective | 4402 (NR) | NR | 88 | 70 | 18 | 18 | 70 |
| Silver et al (2020) (60) | USA | Retrospective | 249 (M=110; F=139) | 59.6 | 6 | NR | NR | NR | NR |
| Stoeckle et al (2020) (61) | USA | Retrospective | 120 (M=96; F=24) | 60.5 (range 56.6-70.0) | 30 | 24 | 2 | 4 | NR |
| Tesoriero et al (2021) (62) | USA | Descriptive | 378248 (M=192646; F=183319) | NR | 2988 | 689 | 207 | 896 | 2092 |
| Virata et al (2020) (63) | USA | Retrospective | 40(M=20; F=20; All PLWH) | NR | 40 | 40 | 0 | 4 | 36 |
| Vizcarra et al (2020) (64) | Spain | Prospective | 269417 (NR) | NR | 51 | 44 | 2 | 6 | 45 |
| Wang et al (2020) (65) | China | Descriptive | 125 (M=71; F=54) | 38.76±13.799 | 1 | 1 | 0 | NR | NR |
| Yang et al (2021) (66) | China | Retrospective | 188 | NR | 3 | NR | NR | NR | NR |
| Yu et al (2020) (67) | China | Retrospective | 142 (M=81; F=61) | 61.9±12.4 | 8 | NR | NR | NR | NR |

M, Male; F, Female; T, Transgender man/woman; SD, Standard Deviation; IQR, Interquartile Range; PLWH, People Living with HIV; NR, Not Reported.

### Prevalence of PLWH co-infected with Sars-CoV-2

Of the 46 studies included, 10 studies reported only PLWH-COVID-19 and were not included in the analysis (30, 33, 40, 48, 54, 63). Another study in which patients were not randomly sampled (61) was also excluded from the analysis to reduce bias. Twelve of these studies were conducted in Europe with 9 and 8 studies from Africa and Europe, respectively. The global pooled prevalence of PLWH diagnosed with COVID-19 was 1% (95% CI = 0.9% -1.1%, p < 0.001) while on the continental level, the pooled prevalence of Europe and USA were 0.3% and 1.2% respectively. Also, 67% (6/9) of the studies from the USA were from states (New York and Georgia) within the top 10 highest HIV infection rates based on recent data (68) which may explain the higher prevalence compared with Europe. The pooled prevalence of studies from Africa was expectedly the highest at 11% (95% CI, 4%-18%) while that of continental Asia was not significant. Further, 67% (6/9) of the studies from Africa are from East and Southern Africa; the region with over half (55%) of the total global HIV infections according to the 2021 estimate (8). The variation in the prevalence of HIV infection in this study is illustrative of the current global epidemiology of HIV whereby more than two third of PLWH are currently in Sub-Saharan Africa (69). Moreover, the overall between-studies heterogeneity was significantly high (I^2^ = 99.7%, p<0.001; Fig. 2a) and this is expected due to the variation in global distribution of HIV infections. Publication bias in the pooled studies was further assessed by computing a funnel plot and Egger’s test which was significant (T(95%CI) = 2.17 (0.39 – 12.18), p=0.04; Fig 2b).

**Fig 2a.**
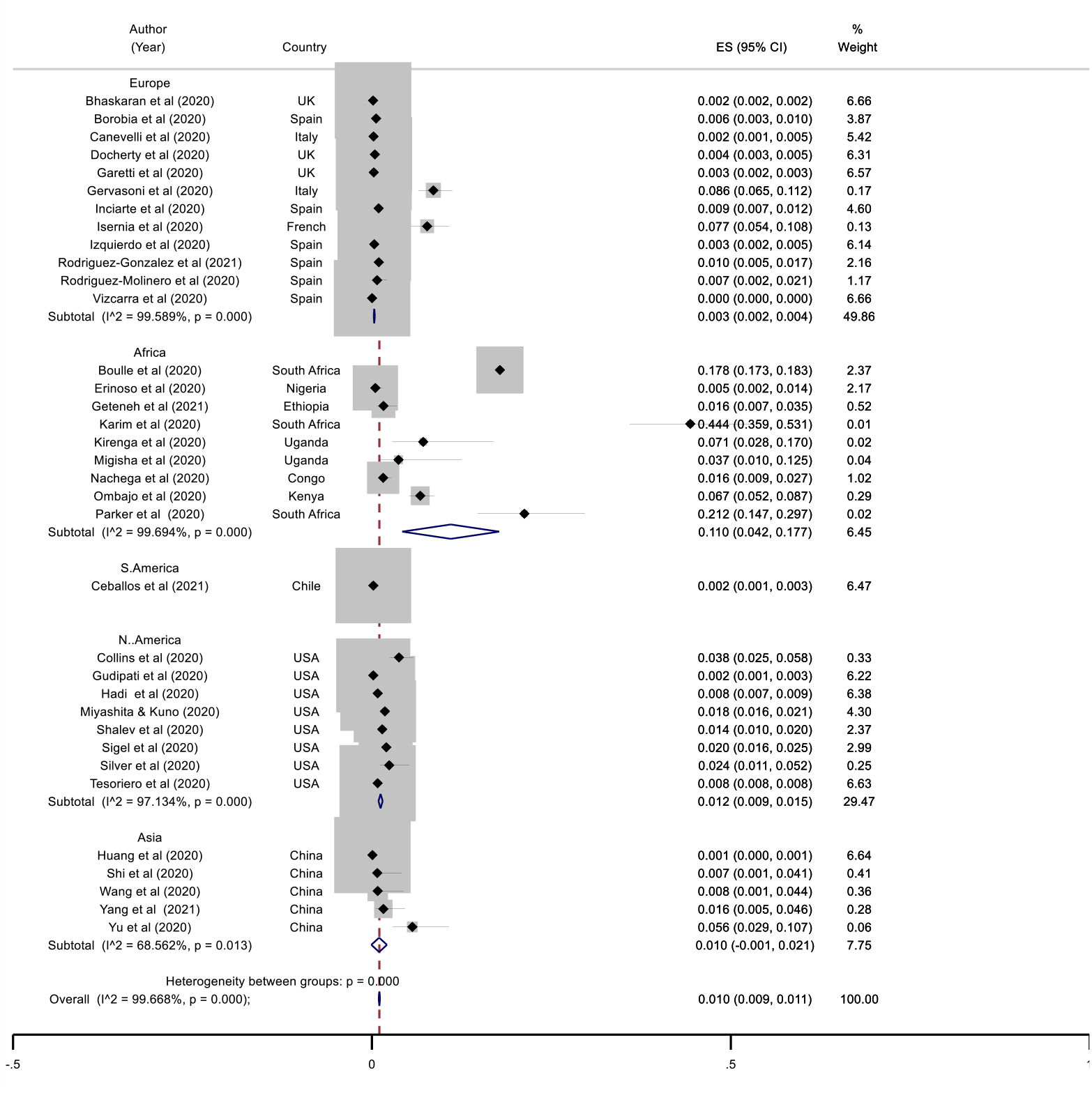
Pooled prevalence of PLWH co-infected with Sars-CoV-2, (ES: Effect size, CI: Confidence Interval).

**Fig 2b.**
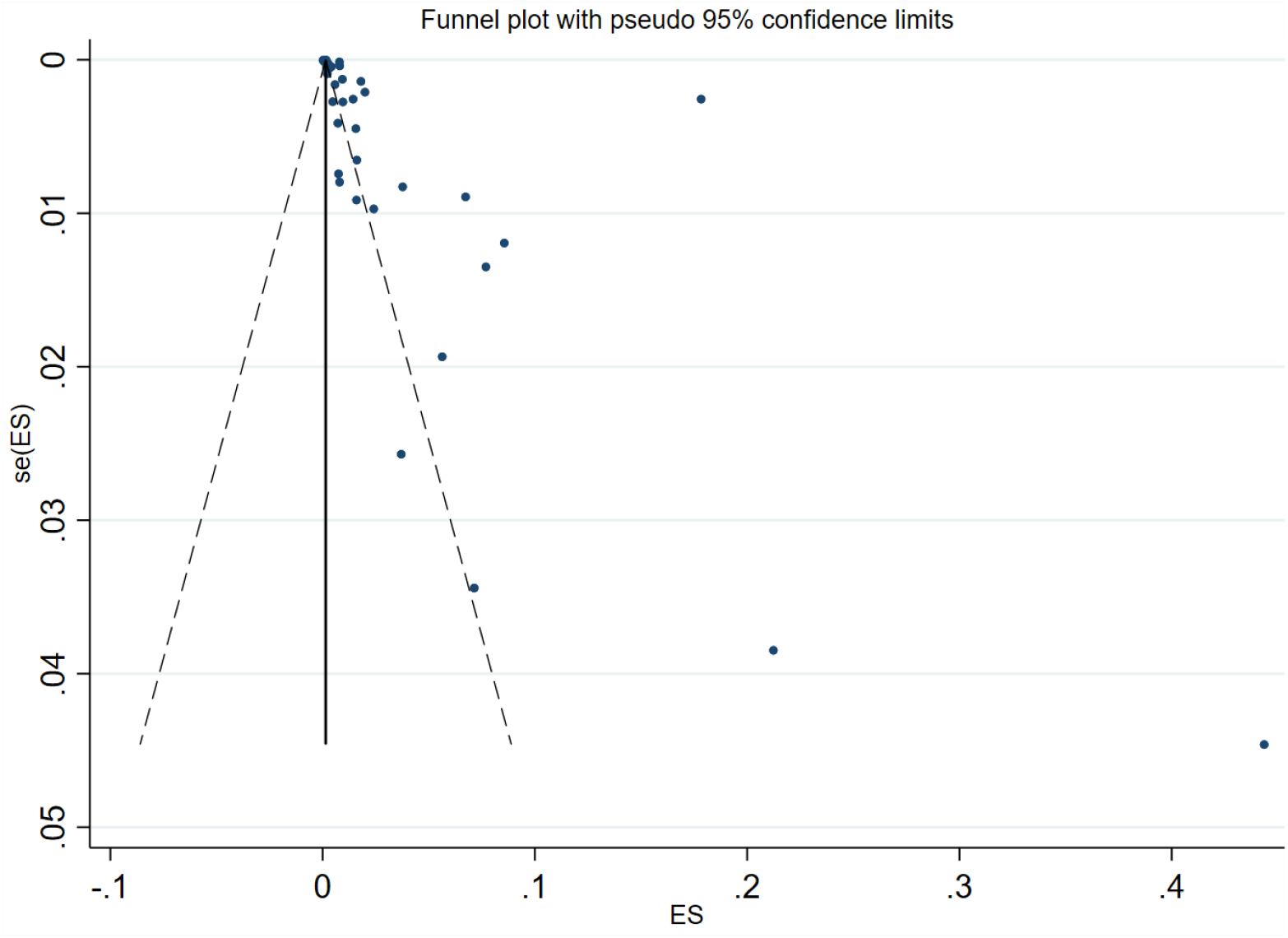
Funnel plot of studies pooled for prevalence of PLWH in COVID-19, (ES: Effect Size, se: standard error).

### Severity of COVID-19 in PLWH

Thirteen studies presented data on severity of COVID-19 in PLWH and non-HIV patients and were computed to determine the risk of COVID-19 severity in PLWH. These studies included a total of 485660 COVID-19 patients of which 7798 (1.6%) were PLWH. Overall, 5, 4 and 2 of the pooled studies were conducted in Africa, the USA and Europe, respectively. The pooled global risk ratio was not significant and shows that PLWH may not be at risk of developing severe COVID-19 (RR (95% CI) = 1.21(0.99 – 1.48); p = 0.477; Fig 3a). This result holds true for both Europe and USA both of which are associated with better prevention and management of HIV. However, the risk for severe COVID-19 in PLWH from Africa was found to increase by 14% (RR (95%CI) = 1.14 (1.05 – 1.24). Also, while the overall heterogeneity was significantly high (85%, p < 0.001), there was no between study variation in the studies from Africa (I^2^ = 0%, p = 0.43). Indeed, funnel plot and Egger’s test showed no publication bias in all included studies (T(95%CI) = -1.32(−3.02 -0.75), p = 0.21; Fig 3b).

**Fig 3a.**
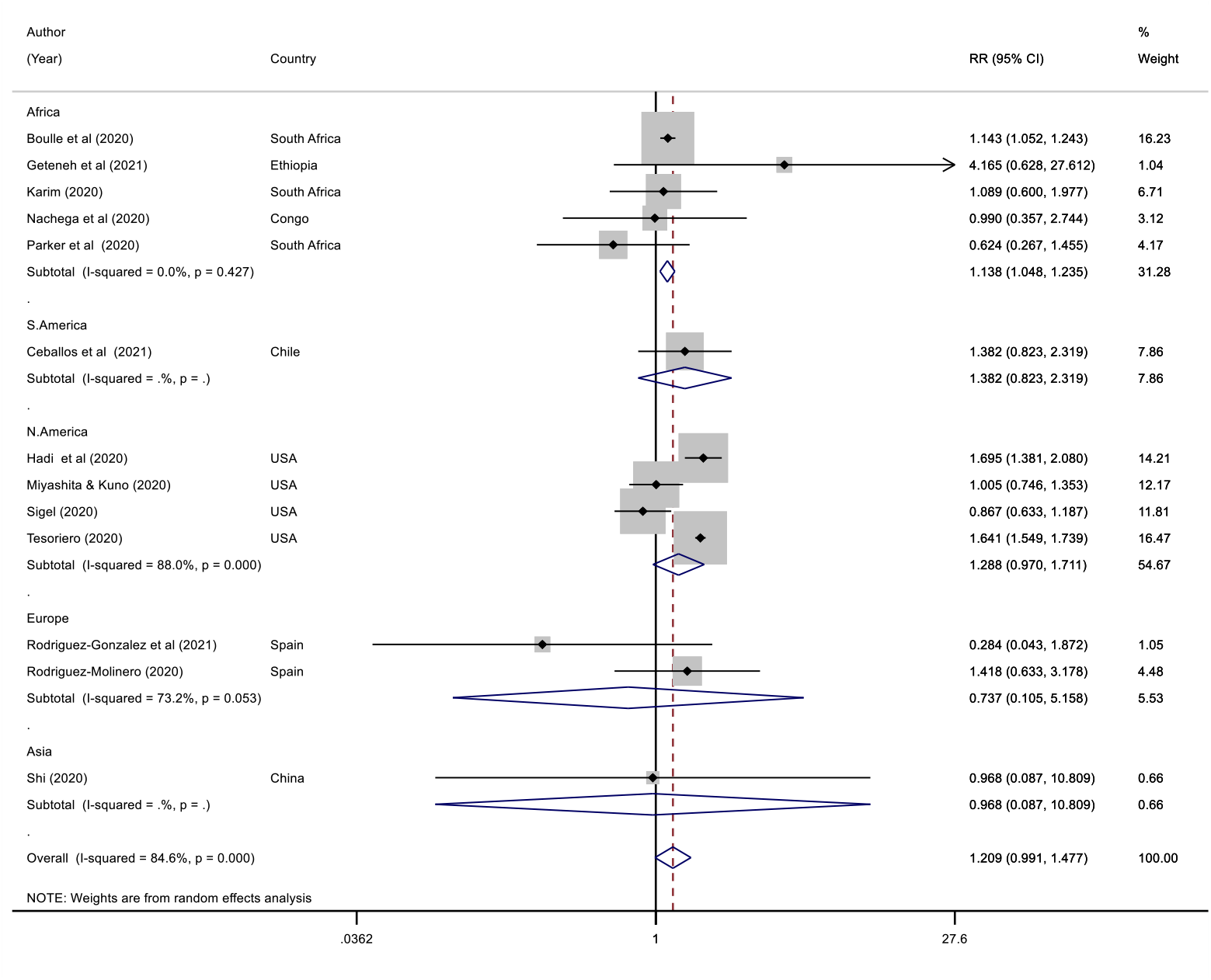
Forest plot of studies pooled for the risk of severe COVID-19 in PLWH, (RR: Risk Ratio, CI: Confidence Interval).

**Fig 3b.**
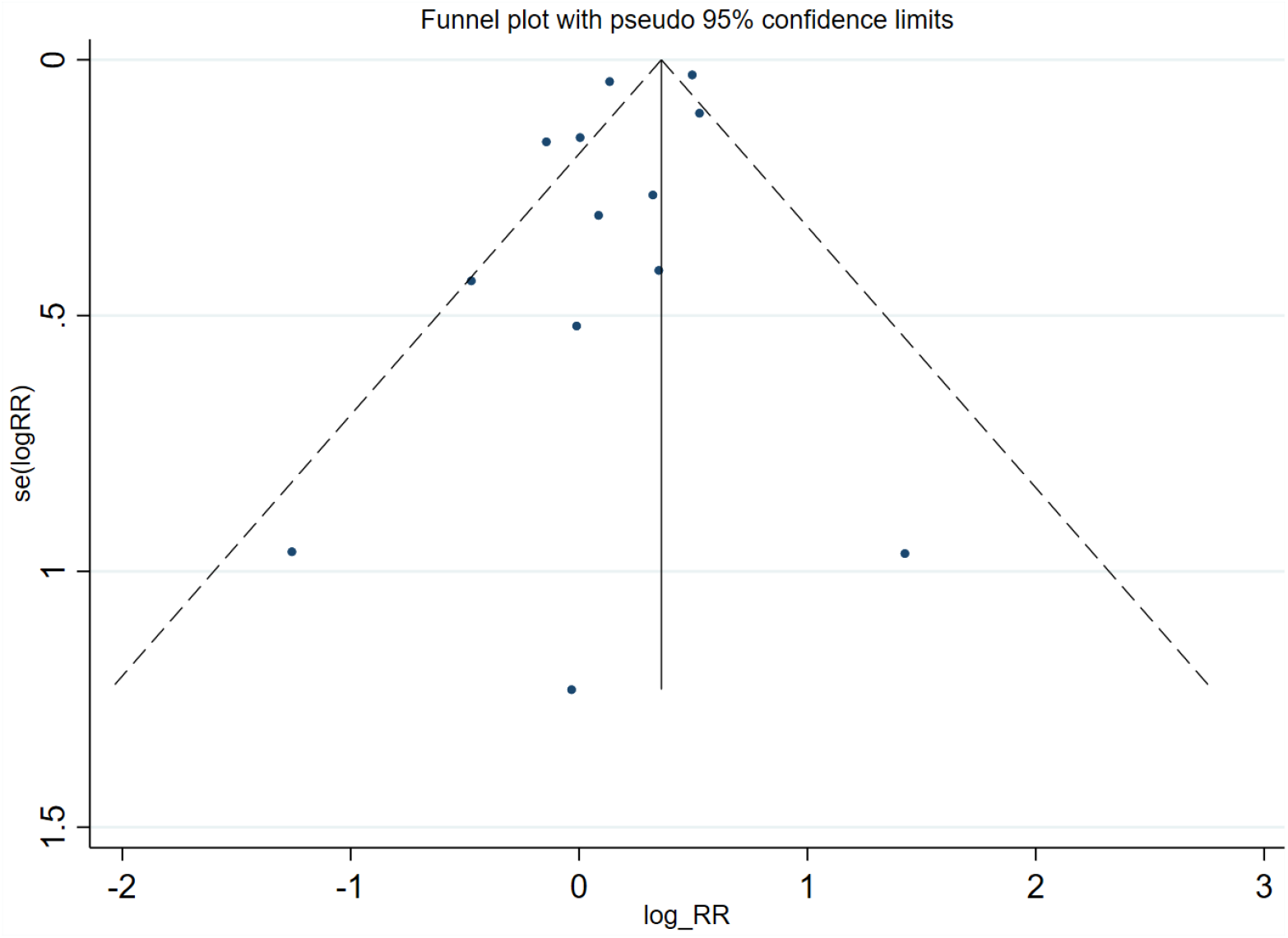
Funnel plot of studies pooled for the risk of severe COVID-19 in PLWH, (RR: Risk Ratio, se: standard error, log: natural logarithm).

### Mortality of PLWH co-infected with SARS-CoV-2

A total of 20 studies were included in the assessment of the risk of mortality from COVID-19 in PLWH. The 20 studies have 17900136 COVID-19 patients including 35582 (0.2%) PLWH. Most of the studies pooled were from Europe (40%, 8/20) with 5 each conducted in Africa and the USA. The analysis for the risk of death in PLWH co-infected with SARS-CoV-2 showed PLWH have 52% increased risk of death from COVID-19 on the global level (RR (95%CI) = 1.52(1.14 – 2.03); Fig 4a). On the regional level, there was no significant increased risk of COVID-19 mortality in PLWH in Africa or Europe. But a 2-fold increase in severity was observed in the USA for the studies included. Despite this difference in regional risk ratios, the computed funnel plot and Egger’s test showed no publication bias in the included studies (T(95%CI) = -0.44 (−1.58 -2.41); Fig 4b).

**Figure 4a.**
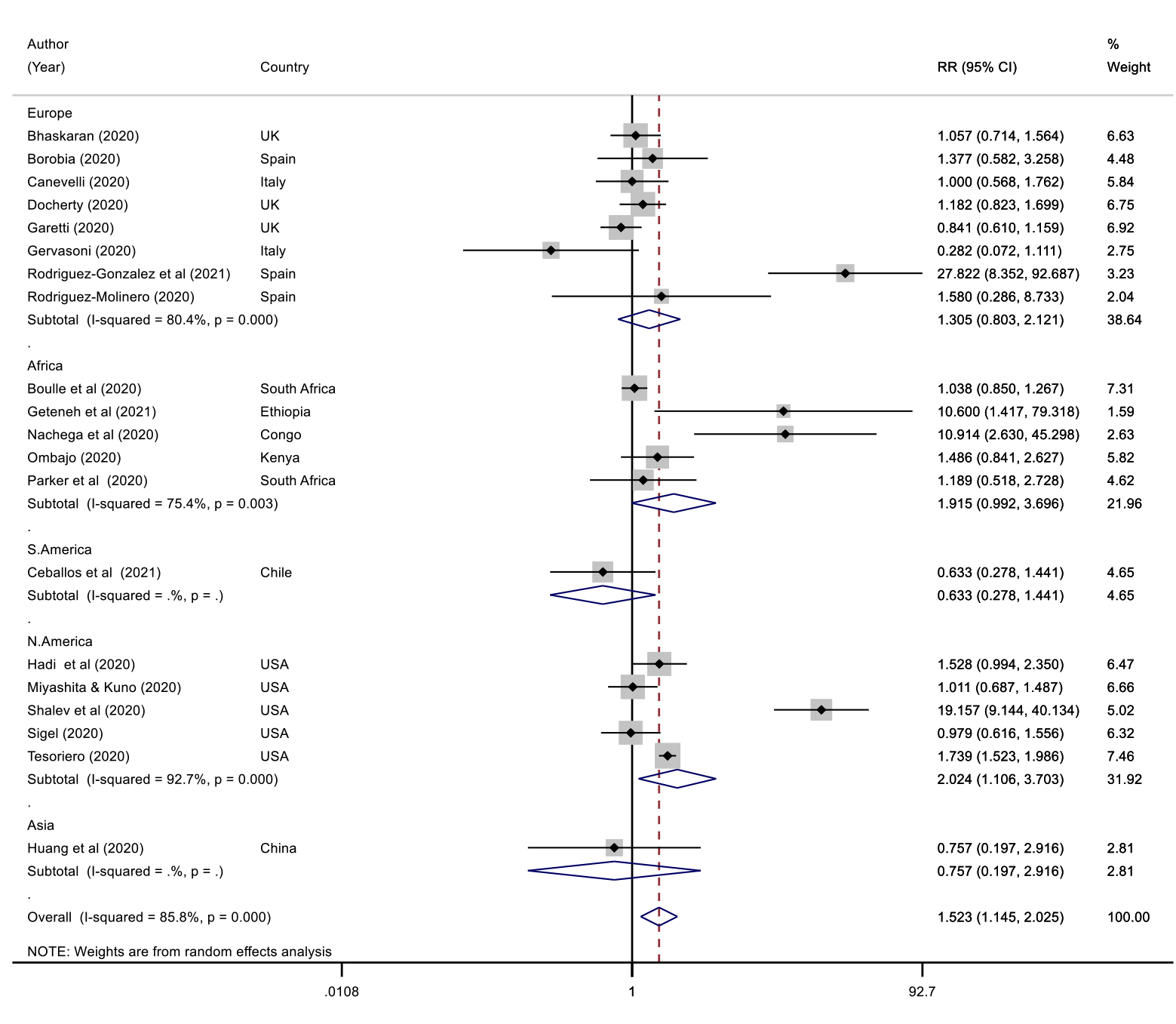
Forest plot for COVID-19 mortality in PLWH, (RR: Risk Ratio, CI: Confidence Interval).

**Figure 4b.**
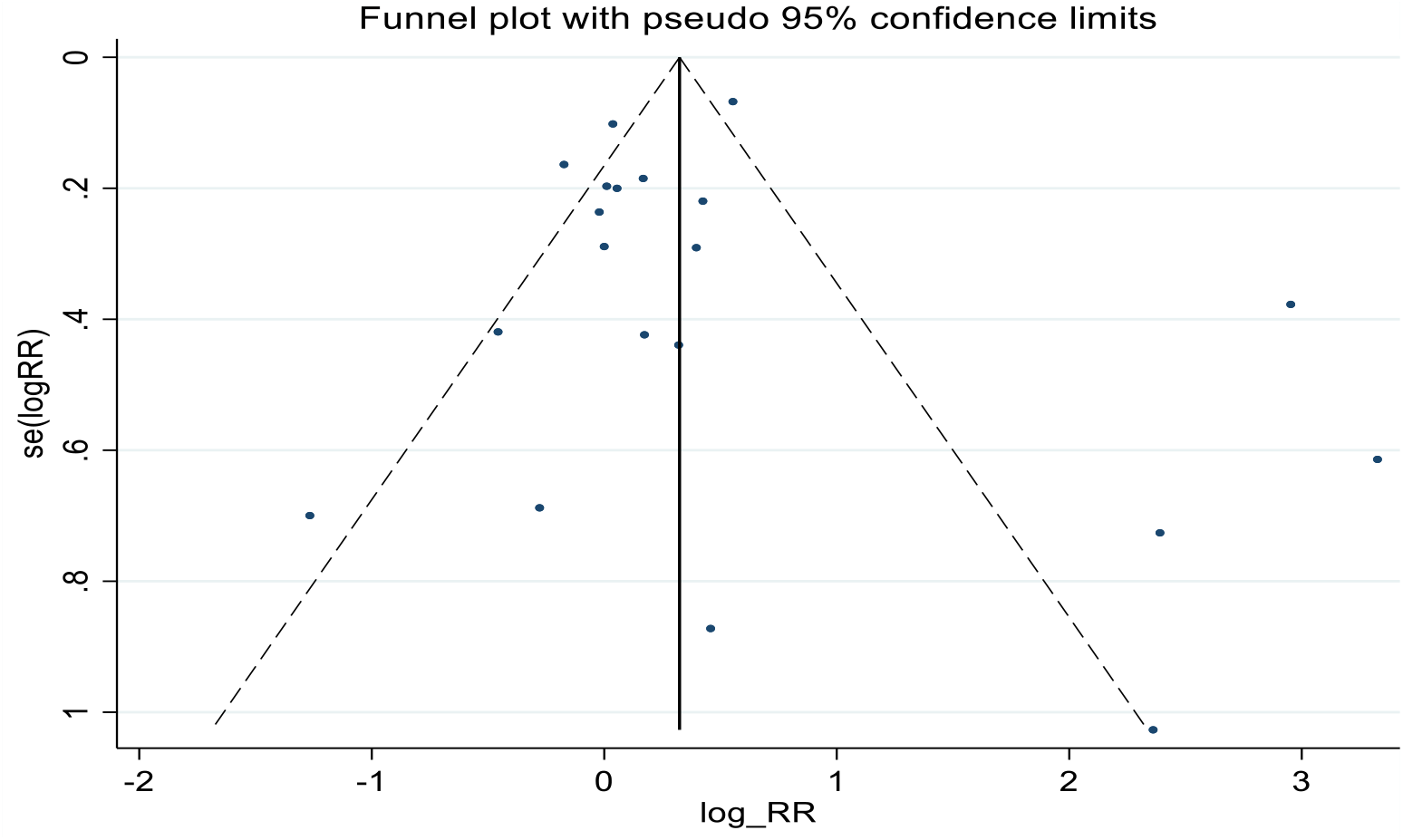
Funnel plot of studies pooled for COVID-19 mortality in PLWH, (RR: Risk Ratio, se: standard error, log: natural logarithm).

## Discussion

This study provides an updated systematic assessment of the prevalence and clinical outcomes of COVID-19 in PLWH on both the global and regional level. To the best of our knowledge, this is a first systematic review and meta-analysis dedicated singly to the understanding of the clinical outcome of PLWH co-infected with SARS-CoV-2 in both global level and within continents with consideration of the variation in the prevalence, prevention, and management of HIV infections. We found a significantly low global prevalence of PLWH in COVID-19 cases (1%) compared to other comorbidities such as COPD, cardiovascular, chronic liver, and kidney disease (2, 3, 70, 71). This is consistent with previous reports which estimate the prevalence of HIV coinfection at 1-2% of COVID-19 patients either admitted to the hospital or in the general population (72). Further, while the proportion was below 2% in Europe, North America and Asia, the prevalence of HIV infection in COVID-19 cases was high in Africa (11%). This is reflective of the global epidemiology of PLWH whereby more than half of the global cases are located within continental Africa. Interestingly, 67% of the studies from Africa are also within the East and Southern Africa region which accounts for over 54% of the global HIV cases (73). Our finding is consistent with earlier systematic reviews which showed similar global prevalence of PLWH in COVID-19 cases (19, 74).

Our result also shows that PLWH may not be at higher risk of severe COVID-19 defined by admission to intensive care units or need for mechanical ventilation on the global level. Interestingly, this lack of association between HIV infection and COVID-19 severity hold true in Europe and the United States, but not in Africa. We found a 14% increase in risk of severity of COVID-19 for PLWH in Africa. Moreover, 60% (3/5) of the studies analysed for the risk of severe COVID-19 in Africa were conducted in South Africa and all studies originated from sub-Saharan Africa: a region associated with high HIV infection rate and poorer antiretroviral treatment (ART) availability (73). Further, we found a 52% increase in the risk of death from COVID-19 in PLWH on the global level. However, only the North American (United States, USA) continent showed a significant risk of mortality (2-fold) among the regions computed. The 2-fold increased risk found in the United States was attributable to the study by Shalev et al included in the meta-analysis. Interestingly, this significant risk ratio disappeared on the continental and the global risk of mortality on the global level fell to 30% when the Shalev et al study was removed (fig. S2a). However, the Egger’s test showed no significant small study effect when the study was included (fig. 4 vs fig. S2). It should be noted also that the average age of PLWH in the Shalev study is 60.7 years, 71% of whom have more than one COVID-19 outcome-defining comorbidities. Moreover, most studies (6/9, 67%) within the USA were conducted in Georgia and New York, both of which are on the top ten states with highest HIV infections and hardest hit by COVID-19 pandemic (1, 75). Importantly, our findings corroborate some previous reports on the potential risk of severe clinical course of COVID-19 in PLWH. Specifically, various meta-analysis conducted on the difference in risk of severe COVID-19 between HIV-positive and HIV-negative patients with SARS-CoV-2 infection whereby risk of severe COVID-19 and mortality were found to be associated with HIV status (76-78). However, other reports have been conflicting with no difference in risk of severe COVID-19 or death between HIV-positive and HIV-negative patients (79-81) with one report proposing a protective effect of HIV infection against COVI-19 (82). Further, Liang et al reported that HIV infection was not related to poorer outcome of COVID-19 and concluded that any risk observed in HIV-SARS-CoV-2 co-infection may be related to presence of concomitant comorbidities which may be common in patients with undiagnosed or untreated HIV infection (20). The observed risk of severe illness (Africa) and death (globally) from COVID-19 in these studies may be attributed to the interplay between several factors. Firstly, the availability of effective HIV management tools in the developed countries means that PLWH now live longer in these regions (83). Increased age is associated with senescence of the natural immune system which may combine with other immune-dampening features of chronic, untreated HIV infection to increase the risk of severity and death from COVID-19. Also, PLWH especially those with undiagnosed or uncontrolled infections, low CD4 count, opportunistic infections and high viral load may present with severe COVID-19 and are at higher risk of death (84). Aside from CD4 and CD8 T-cells activation, effective and early immunoglobin G (IgG) generation results in effective SARS-CoV-2 clearance and improves clinical outcomes (16, 85). However, uncontrolled HIV replication, may trigger increased CD8 T cells activation, inflammation, T cells exhaustion and dysfunction in B cells activities (86, 87). The combined breakdown of B and T cells functions resulting from natural immune system exhaustion may not only result in poorer COVID-19 outcomes but also compromise the efficacy of vaccines in PLWH. Indeed, the response to and efficacies of various vaccines including hepatitis B, pneumococcal, influenza have been shown to be diminished in PLWH and repeated or modified vaccine administration have been recommended (88-90). However, evidence on COVID-19 vaccine efficacy in PLWH is scarce and vaccination of HIV-positive and negative remain similar. Effective ART can attenuate most of the immune dysregulation resulting from uncontrolled HIV infection and replication and is highly recommended. However, undiagnosed HIV infection and low uptake of ART both of which are prevalent in Africa may predispose to poorer COVID-19 clinical outcomes (8).

Moreover, the prevention (sensitisation and pre-exposure prophylaxis), diagnosis and management (ART) of HIV and other chronic diseases have been affected by the global shift in medical resources to contain the COVD-19 pandemic (16). This shift has been suggested to be a contributary factor to the susceptibility of affected groups to severe COVD-19 and death (91-93). Expectedly, the disruption to healthcare systems especially HIV clinics and the downstream effect have been relatively worse in developing countries possibly resulting in worse outcome for PLWH co-infected with COVID-19 (16, 94). However, more data will be needed to establish the extent of these disruptions in regions already behind in the fight against HIV.

Put together, our result show that while the risk of severity illness and death due to COVID-19 increases respectively in Africa and globally, the mechanistic link between HIV infection and the clinical course of COVID-19 may be more complex than previously thought. Firstly, the regional aggregation performed in this study showed that the prevalence of PLWH in COVID-19 cases can be best translated in the background of current global epidemiology of HIV infections. Indeed, the variation introduced by the differences in regional HIV infection rates makes estimation of global prevalence of HIV-SARS-COV-2 coinfection less reliable if not controlled for regional prevalence of HIV. Secondly, there are complex, hardly resolvable confounders when assessing the relationship between HIV infection and COVID-19 outcome including age, sex, treatment with ART, race, region, immune state of patient, number and forms of comorbidities, duration of comorbidities among other factors. Indeed, Bhaskaran et al (95) controlled for age, sex, ethnicity, comorbidities, and time in a population of COVID-19 patients within the United Kingdom. However, the regional difference in prevalence, prevention techniques and clinical management of both HIV and COVID-19 as well as various social-economic factors mean that their finding may not represent the true link or reflect the situation outside the United Kingdom.

This study has several limitations. Firstly, some of the included studies are case series reporting only the PLWH co-infected with SARS-CoV-2. Such studies were excluded from the prevalence analysis, and this may explain the high heterogeneity in the results. However, the random effect model was used to account for the variations in experimental design. Secondly, most studies did not report the distributions of comorbidities, race, age, CD4 and CD8 counts, duration of HIV infection, ART use among other confounders in the studied groups. Thus, we could not control for these parameters in this study. Also, most studies did not report the clinical outcomes (death and severity) of COVID-19 in both PLWH and patients who are HIV-negative and could not be included in the relative risk computation. However, the Egger’s test and funnel plots show that there is no publication bias in the analysed data.

Our findings have several clinical and research implications. First, it further widens the body of evidence by including more recent studies to report that PLWH may be at an increased risk of severe COVID-19 and death and which regions may be mostly at risk. Secondly, we show that the risk of severe COVID-19 and death in PLWH varies between continents and may reflect a complex interplay of concomitant contributory factors which may need to be controlled for to better understand the direct or total effect of HIV infection on COVID-19 outcome. Also, the prevalence of HIV-SARS-CoV-2 coinfection is best interpreted in the context of the varied global epidemiology of HIV infection in various regions of the world. Finally, considering the complex effect of HIV infection on the host immune system as well as the dependence of vaccine efficacy on immune response, future studies should assess COVID-19 vaccine pharmacokinetics in HIV-positive patients to decide whether PLWH coinfected with SARS-CoV-2 may benefit from certain types of vaccines, prioritisation, or repeated inoculations.

## Data Availability

All data relating to the report is contained within the manuscript. We are ahppy to provide further information if required.

