## Supplemental materials for "Global and regional prevalence and outcomes of COVID-19 in people living with HIV: A cutting-edge systematic review and meta-analysis"

Table S1 - Quality Assessment.

| Author (publication year) | SELECTION | | | | ASCERTAINMENT | | OUTCOME | | | OVERALL SCORE (≥5 Stars = Lower Risk of Bias) |
|---|---|---|---|---|---|---|---|---|---|---|
| | Adequate sample size (>=100) | Sample representative of population (Multicentre) | Both groups drawn from same community | Randomness in recruitment | WHO-standard diagnosis (RT-PCR) | HIV Comorbidity confirmation (EMR/Test) | Outcome Reported Adequately (EMR) | Follow up duration adequate (>= 2 weeks (95)) | All subjects accounted for | |
| Bhaskaran et al (2020) (22) | * | * | * | * | - | * | * | * | - | 7 |
| Biagio et al (2020) (23) | * | * | * | - | * | * | * | - | * | 7 |
| Borobia et al (2020) (24) | * | - | * | - | - | * | * | - | * | 6 |
| Boulle et al (2020) (25) | * | * | * | * | * | * | * | * | - | 8 |
| Byrd et al (2020) (26) | - | - | * | - | * | * | * | * | * | 7 |
| Canevelli (2020) (27) | * | * | * | * | * | * | * | * | * | 9 |
| Ceballos et al (2021) (28) | * | * | * | * | * | * | * | * | * | 9 |
| Collins et al (2020) (96) | * | * | * | * | * | * | * | - | * | 7 |
| Del Amo et al (2020) (30) | * | * | * | - | * | * | * | * | * | 8 |
| Docherty et al (2020) (31) | * | * | * | * | - | * | * | * | * | 8 |
| Erinoso et al (2020) (97) | * | * | * | * | * | * | * | * | * | 9 |
| Etienne et al (2020) (33) | - | - | - | - | - | * | * | * | * | 4 |
| Garetti et al (2020) (34) | * | * | * | * | * | * | * | * | * | 9 |
| Gervasoni et al (2020) (35) | - | - | * | - | - | * | * | * | - | 4 |
| Geteneh et al (2021) (36) | * | - | * | * | * | * | * | - | - | 6 |
| Gudipati et al (2020) (98) | * | - | * | - | - | * | * | - | - | 4 |
| Hadi et al (2020) (38) | * | - | * | - | - | * | * | * | - | 5 |

| | | | | | | | | | | |
|---|---|---|---|---|---|---|---|---|---|---|
| Harter et al (2020) (39) | - | - | - | - | * | * | * | * | - | 4 |
| Ho et al (2020) (40) | - | * | * | - | - | * | * | - | * | 5 |
| Huang et al (2020) (99) | * | * | * | * | * | * | * | - | * | 8 |
| Inciarte et al (2020) (100) | * | - | * | * | * | * | * | - | * | 7 |
| Isernia et al (2020) (101) | - | - | - | - | - | * | * | * | * | 4 |
| Izquierdo et al (2020) (102) | * | * | * | * | * | * | * | - | * | 8 |
| Karim et al (2020) (45) | * | - | * | * | * | * | * | * | * | 8 |
| Kirenga et al (2020) (46) | - | - | * | * | * | * | * | * | * | 7 |
| Liu (2020) (47) | - | - | - | - | * | * | * | * | * | 5 |
| Maggiolo et al (2020) (48) | - | - | - | - | * | * | * | * | - | 5 |
| Migisha et al (2020) (49) | - | * | * | - | - | * | * | - | * | 4 |
| Miyashita & Kuno (2020) (50) | * | * | * | * | * | * | * | - | * | 8 |
| Nachega et al (2020) (51) | * | * | * | * | * | * | * | * | * | 9 |
| Ombajo et al (2020) (52) | * | * | * | * | * | * | * | - | * | 8 |
| Parker et al (2020) (53) | * | - | * | * | * | * | * | * | * | 8 |
| Pujari et al (2021) (54) | - | - | * | - | * | * | * | - | * | 5 |
| Rodriguez-Gonzalez et al (2021) (55) | * | - | * | * | * | * | * | * | * | 8 |
| Rodriguez-Molinero et al (2020) (56) | * | * | * | * | * | * | * | - | * | 8 |
| Shalev et al (2020) (57) | - | - | * | * | * | * | * | - | * | 6 |
| Shi et al (2020) (103) | * | - | * | * | * | * | * | * | * | 8 |
| Sigel et al (2020) (59) | - | * | * | * | - | * | * | * | * | 7 |
| Silver et al (2020) (60) | * | - | * | * | * | * | * | * | * | 8 |
| Stoeckle et al (2020) (61) | * | * | - | - | - | * | * | * | * | 6 |
| Tesoriero et al (2020) (62) | * | * | * | - | * | * | * | - | * | 7 |

| | | | | | | | | | | |
|---|---|---|---|---|---|---|---|---|---|---|
| Virata et al (2020) (63) | * | - | * | - | - | * | * | * | * | 6 |
| Vizcarra et al (2020) (64) | * | - | * | - | * | * | * | * | * | 7 |
| Wang et al (2020) (65) | * | - | * | * | * | * | * | * | * | 8 |
| Yang et al (2021) (66) | * | - | * | * | * | * | * | * | - | 7 |
| Yu et al (2020) (104) | * | - | * | * | - | * | * | - | * | 6 |

PCR, Polymerase Chain Reaction; EMR, Electronic Medical Record.

Database: Embase Classic+Embase <1947 to 2021 June 22>
Search Strategy:
--------------------------------------------------------------------------------
1 exp COVID-19/ (123940)
2 exp Severe acute respiratory syndrome coronavirus 2/ (34233)
3 1 or 2 (128690)
4 exp Human immunodeficiency virus/ (204532)
5 3 and 4 (819)
6 limit 5 to (full text and human and english language) (82)

***************************

Database: Embase Classic+Embase <1947 to 2021 January 10>
Search Strategy:
--------------------------------------------------------------------------------
1 Coronavirus Infections/ or COVID-19.mp. (65781)
2 2019 novel coronavirus.mp. (1305)
3 2019 coronavirus.mp. (344)
4 2019-nCoV.mp. (1337)
5 SARS-nCoV 2.mp. [mp=title, abstract, heading word, drug trade name, original title, device manufacturer, drug manufacturer, device trade name, keyword, floating subheading word, candidate term word] (14)
6 SARS-CoV-2.mp. (22727)
7 1 or 2 or 3 or 4 or 5 or 6 (70423)
8 clinical characteristic*.mp. [mp=title, abstract, heading word, drug trade name, original title, device manufacturer, drug manufacturer, device trade name, keyword, floating subheading word, candidate term word] (123413)
9 clinical characteristics.mp. (121781)
10 clinical feature*.mp. [mp=title, abstract, heading word, drug trade name, original title, device manufacturer, drug manufacturer, device trade name, keyword, floating subheading word, candidate term word] (771891)
11 clinical features.mp. (164456)
12 8 or 9 or 10 or 11 (854476)
13 7 and 12 (3667)
14 limit 13 to (english language and full text) (520)

***************************

Database: Ovid MEDLINE(R) ALL <1946 to June 22, 2021>
Search Strategy:
--------------------------------------------------------------------------------
1 exp COVID-19/ (86546)
2 exp SARS-CoV-2/ (67048)
3 exp Coronavirus/ or exp Coronavirus Infections/ (103305)
4 1 or 2 or 3 (103305)
5 *HIV/ or HIV Infections/ or *HIV-2/ or *HIV-1/ (239073)
6 exp Acquired Immunodeficiency Syndrome/ (76801)
7 5 or 6 (294497)
8 4 and 7 (743)
9 limit 8 to (english language and full text and humans) (142)
10 limit 9 to "review articles" (23)
11 9 not 10 (119)

***************************

**Figure S1** – Search Strategy

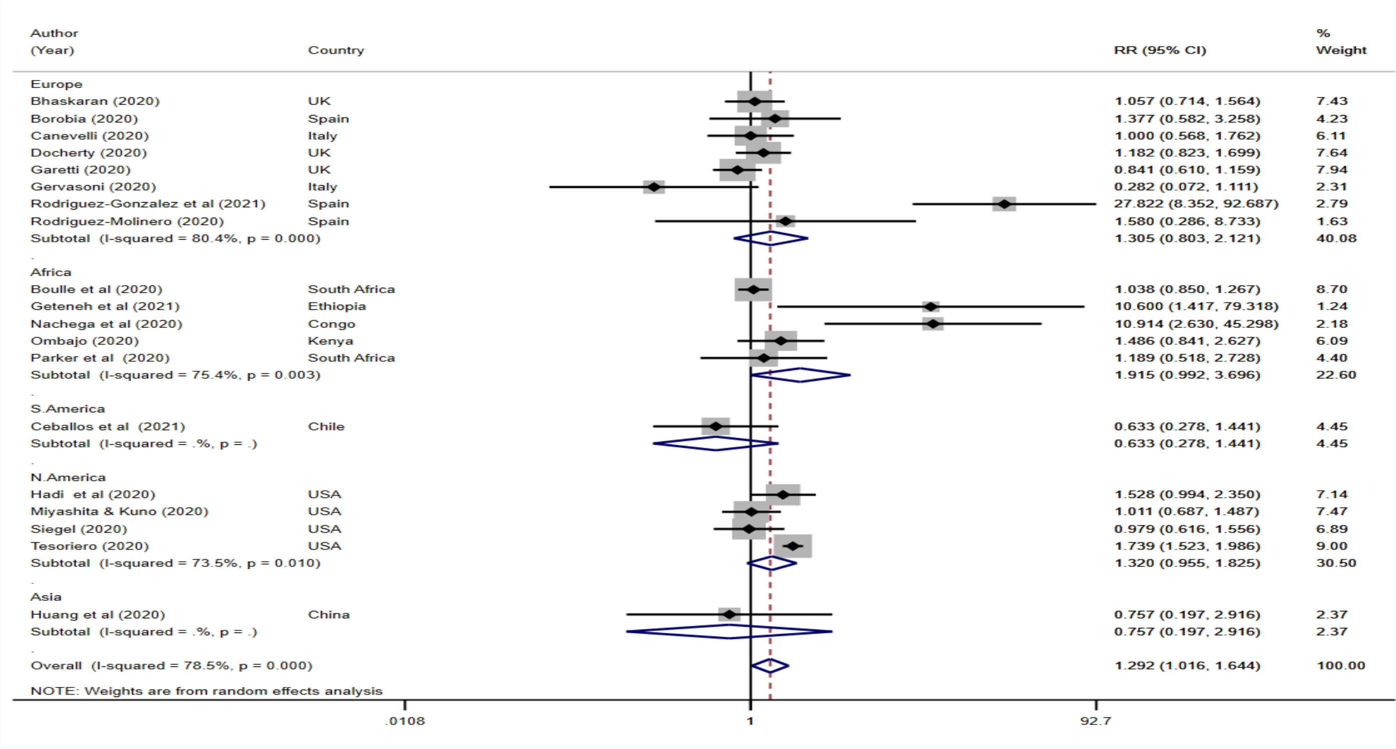


**Figure S2a** - Forest plot for COVID-19 mortality in PLWH without the study by Shalev et al (2020) *(57)*, (ES: Effect size, CI: Confidence Interval).

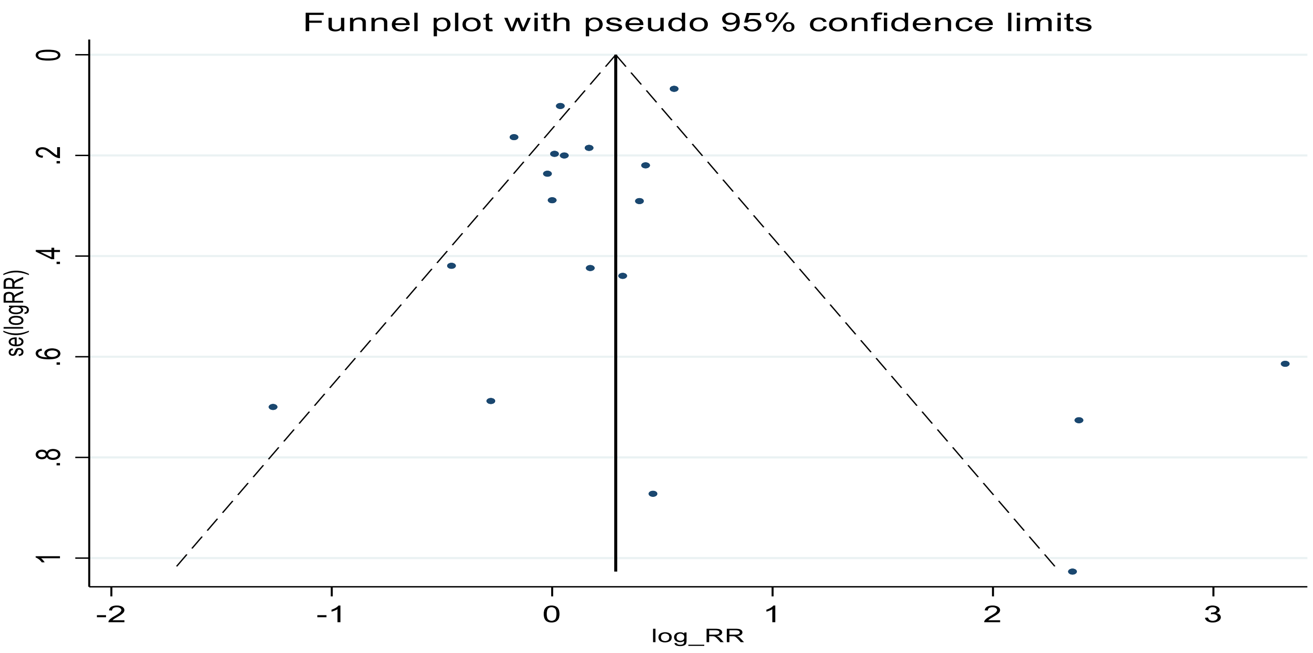


**Figure S2b** - Funnel plot of studies pooled for COVID-19 mortality in PLWH without the study by Shalev et al (2020) *(57)*, (RR: Risk Ratio, se: standard error, log: natural logarithm).
